# Barriers and enablers to participation in a proposed online lifestyle intervention for older adults with age-related macular degeneration

**DOI:** 10.1101/2023.05.24.23290417

**Authors:** Richard Kha, Qingyun Wen, Nicholas Bender, Charlotte Jones, Bamini Gopinath, Rona Macniven, Diana Tang

**Author notes:** This project is funded by the Macular Disease Foundation Australia. The authors declare no competing interests.

## Abstract

Age-related macular degeneration (AMD) is a blinding condition associated with depression and loneliness. This facilitates unhealthy lifestyle behaviours which drives AMD progression. We developed the first online lifestyle intervention for AMD, called Movement, Interaction and Nutrition for Greater Lifestyles in the Elderly (MINGLE) to promote positive lifestyle changes, reduce loneliness and depression. This qualitative study explored enablers and barriers to participation in MINGLE for older Australians with AMD. Thirty-one participants with AMD were interviewed using a semi-structured in-depth approach. Thematic analysis revealed nine themes. Enablers to participation were: socialising and learning about AMD, motivation to improve health, program accessibility and structure. Barriers were: lack of time, unfamiliarity with technology, limited knowledge regarding holistic interventions, vision-related issues, mobility and negative perception of group interactions. Multiple factors influence the participation of AMD patients in MINGLE and these must be considered when developing and implementing the MINGLE program to maximise participation.

## Introduction

Age-related macular degeneration (AMD) is the leading cause of irreversible blindness among adults in developed countries (Mitchell et al., 2018). Globally, 196 million individuals are living with AMD and this number is projected to increase to 288 million by 2040 (Wong et al., 2014) due to the increase in life expectancy, making AMD a significant challenge for healthcare systems (Wong et al., 2014). Approximately 85% of all AMD patients are affected by early-stage and late-stage dry AMD, while 15% are affected by late-stage wet AMD (Cabral de Guimaraes et al., 2022). Although pharmacological treatments exist for wet AMD, they do not exist for the more prevalent early- and dry-AMD (Hadziahmetovic & Malek, 2020).

AMD is a chronic degenerative condition that causes irreversible central vision loss. Affected individuals may find it difficult to recognise faces and perform everyday tasks such as driving and reading (Choudhury et al., 2016), which has profound negative impacts on overall wellbeing. AMD is a significant risk factor for depression, presumably due to the loss of ability to partake in valued activities and reduced function (Casten & Rovner, 2013). Research shows that the prevalence of depression among people with AMD ranges from 15.7-44% (Dawson et al., 2014; Heesterbeek et al., 2017). Older adults with visual impairment are also twice as likely to experience loneliness compared to those with normal vision (Alma et al., 2011). This is supported by findings from a Norwegian study where the prevalence of moderate and severe loneliness in older adults was significantly greater in visually impaired adults compared to the general population (Audun Brunes et al., 2019).

The follow-on effects of depression and loneliness include unhealthy lifestyle behaviours including sedentary behaviours and unhealthy diets (Loprinzi et al., 2015). These behaviours are associated with an increased risk of AMD development and progression (Schrempft et al., 2019; Tang et al., 2020). Persistent depression and loneliness also contribute to other serious health outcomes such as cardiovascular disease, frailty and dementia (Schrempft et al., 2019; Sutin et al., 2020).

Holistic lifestyle interventions involving education and group support components can reduce depression and loneliness in older people (Senra et al., 2019). Studies have shown that psychosocial treatments such as self-management programs, group problem-solving sessions and mindfulness activities reduced depression and loneliness among individuals with AMD (Senra et al., 2019). Additionally, non-pharmacological lifestyle changes such as smoking cessation, adoption of a Mediterranean diet and increasing physical activity have been shown to reduce onset and progression of AMD (Carneiro & Andrade, 2017; Di Carlo & Augustin, 2021). The increasing evidence supporting the benefits of lifestyle changes for adults with AMD over the past decade has brought greater attention to the need for holistic support programs which encompass emotional, practical, informational and peer support for visually impaired adults, especially those with AMD (Barrow et al., 2018; Rabiee et al., 2015).

Despite the need for lifestyle interventions that integrate group social support, physical activity and dietary education for patients with AMD, no such program has been developed yet. Our group has previously developed evidence-based interventions which have addressed different aspects of holistic lifestyle interventions. For example, we previously created the Walk N’ Talk for your Life (WTL) program to target social isolation and loneliness in older adults (Jones et al., 2019). WTL was a 12- and 10-week group program for older adults delivered in-person and over Zoom in Canada and the United Kingdom, respectively (Jones et al., 2019). Socialisation was a key aspect of the WTL program, which included resistance training, balance exercises, group walking and discussion of strategies to improve health (Jones et al., 2019). This program was well received by participants and reduced feelings of loneliness and symptoms of depression by 30% (Jones et al., 2019). A qualitative study was conducted after the WTL program exploring motivators and barriers to help-seeking among participants in the program (Montagliani et al., 2018). A barrier identified in this study included physical barriers such as the lack of access due to geographical location, which inspired a need to explore an online program such as MINGLE (Montagliani et al., 2018).

We also previously developed a telehealth dietary behavioural program for AMD patients which resulted in improved dietary habits and AMD-specific nutrition knowledge. We aim to build upon our prior research by incorporating elements of both the aforementioned WTL (Jones et al., 2019) and dietary behavioural program (Tang et al., 2020) to develop a holistic lifestyle intervention specifically for patients with AMD called Movement, Interaction and Nutrition for Greater Lifestyles in the Elderly (MINGLE). The proposed MINGLE program is a 10-week program which will be delivered online, consisting of weekly sessions involving group exercise, socialisation and education about diet and AMD. Further details of the proposed MINGLE program can be found in the published protocol paper (Tang et al., 2022).

As MINGLE will be the first AMD-related lifestyle intervention to be delivered online, it is important to explore the perspectives of people with AMD to future participation in the proposed MINGLE program. These perspectives will be critical in guiding the development and implementation of the MINGLE program in a future pilot trial to maximise its uptake, adherence and efficacy. This current qualitative study aims to use the theoretical domains framework (Atkins et al., 2017) to investigate the barriers and enablers of people with AMD to future participation in the proposed MINGLE program.

## Methods

### Design and setting

The study design method used for data collection and analysis in this qualitative study was based on principles from classical grounded theory (Glaser & Strauss, 2017). Grounded theory is useful for generating inductively based explanations of psychosocial processes and its explorative nature is valuable for previously unexplored research areas (Glaser & Strauss, 2017). This qualitative study recruited participants with AMD from a private retinal clinic in Sydney, Australia and through a public webinar hosted by the Macular Disease Foundation Australia. Participants provided verbal consent to participate. Inclusion criteria were diagnosis of any form of AMD; ability to speak and understand English; and consent to participating in the study and audio recording.

Semi-structured one-on-one interviews were conducted in-person and over the telephone by two researchers with qualifications in medicine and ophthalmology (RK, QW). Semi-structured interviews were used because of their flexible structure and ability to deeply explore participant perspectives (DeJonckheere & Vaughn, 2019). Informed verbal consent was audio recorded and documented in a Record of Verbal Consent form. Interview questions (Supplementary File 1) explored the participants’ experience with AMD, its impact on their day-to-day life, previous experiences with group programs, and questions relating to the proposed MINGLE program, including duration, frequency and time of the sessions, suitability of the exercises and nutrition topics, as well as thoughts on what to include and/or exclude from the program. The final questions were assigned to elicit information about potential barriers and enablers to future participation in the proposed MINGLE program and strategies to maintain commitment if the participant were to join the MINGLE program. The consolidated criteria for reporting qualitative research checklist guided the reporting of this research (Tong et al., 2007).

### Data collection

The interviews were conducted between November 2021 to January 2022. Interview audios were digitally recorded and transcribed verbatim by an external transcription service. Transcripts were exported as Microsoft Word documents and imported into NVivo 11 (QSR International, Australia). Data collection and analysis occurred simultaneously and ceased once data saturation was reached. Demographic information for each patient was collected from paper records at the clinic. This data included age, sex, AMD type, time since AMD diagnoses, visual acuity in each eye and history of other ocular conditions.

### Data analysis

The data were analysed in NVivo using the theoretical domains framework (TDF) method (Atkins et al., 2017) which involved systematically coding the data into specific topics such as ‘barriers’ and ‘enablers’. TDF has been used in various studies focusing on behaviour patterns of patients, as well as barriers and enablers in uptake of healthcare services (Burgess et al., 2015; Sethu et al., 2022). The data were revisited, coded and themes were discussed multiple times to ensure analytic reflexivity. Two researchers coded the interview transcripts independently before meeting to discuss and gain consensus on prevalent themes along with their illustrative quotes. These were then presented to the principal investigator for feedback and any discrepancies in coding were resolved through discussion with the principal investigator. Inter-rater reliability between data coders was expressed using Cohen’s kappa. The overall Cohen’s kappa value was 0.86 (range for individual themes: 0.58-1.00) which indicated a strong level of agreement between the data coders.

The study was conducted in compliance with the ethical guidelines of the World Medical Association Declaration of Helsinki. It was approved by the Macquarie University Human Research Ethics Committee (HREC) on 10th August 2021 (ID: 10007)

## Results

### Participants

Thirty-one participants (mean age = 79 ± 7.4 years, range = 58 to 89 years) with AMD were recruited from November 2021 to January 2022. One participant was recruited outside the clinic. Participants were mostly female (18/31, 58%) with late-stage AMD (28/31, 90%) in at least one eye. Table 1 shows the demographic data for the participants. Demographic data was unavailable for one participant (Study ID 031). The average duration of the interviews was 10 (range = 6 to 15) minutes and varied in length depending on the extent of the answers.

**Table 1.** Demographic characteristics of the study participants (n = 30)

| Study ID | Sex | AMD type* | Years since AMD diagnosis | Visual Acuity* | History of other eye conditions |
| --- | --- | --- | --- | --- | --- |
| 001 | Male | LE- late AMD (neovascular)<br>RE – early AMD (dry) | LE – 1.5<br>RE – 1.5 | LE – 6/60 -4<br>RE – 6/12 + 2 | Cataract in both eyes (treated) |
| 002 | Male | LE – early AMD (dry)<br>RE – late AMD (neovascular) | LE – 2<br>RE – 6 | LE – 6/6 - 2<br>RE – 6/15 - 2 | Cataract in both eyes (treated) |
| 003 | Male | LE – late AMD (neovascular)<br>RE – early AMD (dry) | LE – 1.5<br>RE – 1.5 | LE – 6/9 -2<br>RE – 6/12 +2 | Cataract in both eyes (treated) |
| 004 | Male | LE – early AMD (dry)<br>RE – late AMD (neovascular) | LE – 3<br>RE – 3 | LE – 6/30 -2<br>RE – 6/48 -2 | Previous right herpetic keratouveitis with cornea scar (treated) |
| 005 | Male | LE – early AMD (dry)<br>RE – late AMD<br>(neovascular) | LE – 1<br>RE – 2 | LE – 6/24-2<br>RE – 6/36-2 | Cataract in left<br>eye (treated) |
| 006 | Male | LE – early AMD (dry)<br>RE – no AMD | LE – 14<br>RE – | LE – 6/12<br>RE – 6/60 | CRVO in right<br>eye |
| 007 | Female | LE – late AMD<br>(neovascular)<br>RE – late AMD<br>(neovascular and<br>geographic atrophy) | LE – 1<br>RE – 3 | LE – 6/15-1<br>RE – 6/60+2 | Cataract in both<br>eyes (treated) |
| 008 | Male | LE – late AMD<br>(neovascular)<br>RE – late AMD<br>(neovascular) | LE – 5<br>RE – 5 | LE – 6/18<br>RE – 3/60 | CRVO in right<br>eye |
| 009 | Male | LE – no AMD<br>RE – late AMD<br>(neovascular) | LE –<br>RE – 4 | LE – 6/12<br>RE – 3/60 | Cataract in left<br>eye, bilateral<br>moderate NPDR |
| 010 | Female | LE – late AMD<br>(neovascular)<br>RE – no AMD | LE – 4<br>RE – 1 | LE – 6/36<br>RE – 6/24 | Cataract in both<br>eyes (treated) |
| 011 | Female | LE – late AMD<br>(neovascular)<br>RE – early AMD (dry) | LE – 2<br>RE – 0.5 | LE – 6/18<br>RE – 6/12 | Bilateral<br>moderate<br>cataracts |
| 012 | Female | LE – no AMD<br>RE – late AMD<br>(neovascular) | LE –<br>RE – 2 | LE – 6/9<br>RE – 6/15 | - |
| 013 | Female | LE – early AMD (dry)<br>RE – late AMD<br>(neovascular) | LE – 2<br>RE – 5 | LE – 6/9<br>RE – 6/15 | Bilateral cataracts<br>(treated) |
| 014 | Male | LE – late AMD<br>(neovascular)<br>RE – late AMD<br>(neovascular with<br>atrophy) | LE – 8<br>RE – 8 | LE – 6/24<br>RE – 6/60 | None |
| 015 | Male | LE – early AMD (dry)<br>RE – late AMD<br>(neovascular with<br>atrophy) | LE – 1<br>RE – 4 | LE – 6/9<br>RE – 6/60 | Glaucoma in both<br>eyes |
| 016 | Female | LE – late AMD<br>(neovascular with | LE – 2<br>RE – 1 | LE – 6/30<br>RE – 6/18 | Pigment epithelial<br>detachment in |
|  |  | atrophy) |  |  | right eye |
|  |  | RE – early AMD (dry) |  |  |  |
| 017 | Female | LE – late AMD<br>(neovascular) | LE – 4<br>RE – 4 | LE – 6/60<br>RE – 6/9 | None |
|  |  | RE – late AMD<br>(neovascular) |  |  |  |
| 018 | Female | LE – late AMD<br>(neovascular) | LE – 6<br>RE – 6 | LE – 6/12<br>RE – 6/60 | Macular hole in<br>right eye |
|  |  | RE – late AMD<br>(neovascular) |  |  |  |
| 019 | Female | LE – early AMD (dry) | LE – 2 | LE – 6/9 | None |
|  |  | RE – late AMD<br>(neovascular) | RE – 5 | RE – 6/24 |  |
| 020 | Female | LE – early AMD (dry) | LE – 6 | LE – 6/15 | Bilateral cataracts |
|  |  | RE – no AMD | RE – 3 | RE – 6/12 | (treated) |
| 021 | Female | LE – late AMD<br>(neovascular) | LE – 5<br>RE – 5 | LE – 6/18<br>RE – 1/60 | None |
|  |  | RE – late AMD<br>(neovascular) |  |  |  |
| 022 | Male | LE – late AMD<br>(neovascular) | LE – 5<br>RE – 5 | LE – 6/15<br>RE – 6/9 | None |
|  |  | RE – late AMD<br>(neovascular) |  |  |  |
| 023 | Female | LE – no AMD | LE – | LE – 6/9 | None |
|  |  | RE – late AMD<br>(neovascular) | RE – 7 | RE – 6/6 |  |
| 024 | Female | LE – late AMD<br>(neovascular) | LE – 2<br>RE – 2 | LE – 6/18<br>RE – 6/18 | CRVO in right<br>eye (treated) |
|  |  | RE – early AMD (dry) |  |  |  |
| 025 | Male | LE – late AMD<br>(neovascular) | LE – 11<br>RE – 16 | LE – 3/60<br>RE – 6/60 | Bilateral<br>glaucoma and<br>cataracts (treated) |
|  |  | RE – late AMD<br>(neovascular) |  |  |  |
| 026 | Female | LE – late AMD<br>(neovascular) | LE – 1<br>RE – 3 | LE – 6/7.5<br>RE – 6/9 | None |
|  |  | RE – late AMD<br>(neovascular) |  |  |  |
| 027 | Male | LE – early AMD (dry) | LE – 3 | LE – 6/30 | Bilateral |
|  |  | RE – no AMD | RE – | RE – 3/60 | cataracts,<br>previous ischaemic optic neuropathy |
| 028 | Female | LE – none | LE – | LE – 6/9 | R cataract |
|  |  | RE – late AMD | RE – 1 | RE – 6/30 | (untreated) |
| 029 | Female | LE – late AMD | LE – 8 | LE – 6/9 | Endophthalmitis |
|  |  | (neovascular) | RE – 8 | RE – 6/9 | in right eye |
|  |  | RE – late AMD |  |  | (treated) |
|  |  | (neovascular) |  |  |  |
| 030 | Female | LE – early AMD (dry) | LE – 3 | LE – 6/15 | None |
|  |  | RE – late AMD | RE – 7 | RE – 3/60 |  |
|  |  | (neovascular) |  |  |  |

\*LE – left eye, RE – right eye

### Themes

A total of nine themes were identified – three for enablers and six barriers. Enablers included opportunity to meet new people and learn more about AMD, motivation to improve health and the MINGLE program’s accessibility and structure. Barriers included time constraints, access to technology, lack of knowledge about lifestyle risk factors, vision and mobility limitations and a negative perception of social group interactions.

#### Enablers to participation

##### Theme 1: The program would give an opportunity to meet new people and learn more about AMD

Many participants, particularly the older ones, wanted to join the program because of the opportunity for social contact. One participant mentioned, “I love to go and mix with people…to do exercise together, you know? That’s something different. And good for the body, too.” Another participant seemed to agree with this:

> “It is important to get that human interaction. At my age, it makes you feel a lot happier if you have someone to talk to…during the COVID, we couldn’t have visitors, you know? And I know how absolutely lonely people were.”

Participants also believed that that social interaction is important for minimising feelings of loneliness, isolation and improving mental health, with a participant stating, “Socialising is always good for your health and mental state. It just removes the boredom, gives you a better personality and makes you happier.”

For other participants, the program was a means of trying new activities, helping and meeting new people. One participant mentioned, “I’m interested in people, what they will do and how they do…personally meeting new people and doing different things that I haven’t done before.” Another participant agreed with this, “I like socialising, helping each other…if somebody needs help, I’d like to help them.”

Majority of participants were concerned about their lack of knowledge regarding AMD due to limited opportunities to ask questions of their ophthalmologists. They hoped that the program would provide opportunities to improve understanding of their AMD.

> “Am I gonna lose my eyesight basically? From the doctor, you don’t get a lot of feedback at all. You’re left hanging in a lot of ways. If you could include a bit of education in the sessions…I think that would be quite encouraging”

Another participant was interested in learning about lifestyle risk factors associated with AMD, of which diet and physical activity have been identified to be, “The exercise and diet thing, it’ll certainly make me come and want to stay.”

Participants also stated that they would like to receive advice on how to adjust to living with AMD from other people who have the condition. One participant quoted, “The most helpful thing would be learning tips and tricks from other people with shared experiences…like learning how to cross the road again when you can no longer see the traffic.”

##### Theme 2: Participants expressed motivation to improve their current health

Another facilitator to participating in the MINGLE program was participant motivation to improve their health and minimise AMD progression. This was highlighted by an elderly participant, “The motivation is your own condition and you want to improve that, not just for yourself but also for other people if possible.” Another participant emphasised the importance of maintaining their vision, “It’s a pretty serious business when it comes to your eyes. Nobody wants to go blind.”

Some participants mentioned exercise as a reason to join the program as it helps maintain their mental and physical health. One person noted, “I think exercise will improve the mental health and strength. This makes you strong and healthy. We need that every day.” This was echoed by another participant who indicated the significance of exercise in their daily life, “If I don’t exercise every morning, I won’t be able to do whatever I want to do.”

Other participants reported that having group exercise in the program was a motivator as they normally would not do exercise when alone. A participant mentioned, “Exercise has never been interesting to me…unless you do it as part of a group, it becomes rather boring and isolating.”

Several participants also highlighted dietary education as a reason to participate in the program. One participant recognised adverse effects of having a bad diet, “People just don’t know…You can’t be eating junk for years and years and expect to have a good life.” Another participant felt that dietary education and exercise would be beneficial for older adults in general, “Exercise and diet, they’re good for people of our age and mostly the people who have AMD are in their 70s and 80s. These would be ideal for them.” This was echoed by another participant, “I think the dietary education is a brilliant idea. Even spending time learning how to read food labels would be great.”

One participant saw the program as an opportunity to help others who are also affected by AMD, “Appealing to the sense of helping future people, because five years down the track, it could be somebody else they know and if they can help them, then all good.”

##### Theme 3: Program accessibility and structure

Other enablers to participation mentioned by our participants included technology, the lack of cost to participate and having a structured program with goals.

Although technology was a barrier for some participants, it was also an enabler for others. Some participants noted difficulty with driving which affected their ability to participate in group programs as they were unable to travel to the required destination. One participant indicated that they rarely drive anymore, “Unless I drive with familiar conditions or in fine weather, I avoid driving.” Another participant mentioned that they would typically have to rely on other people to access face-to-face group programs, “It’s very hard for me to get anywhere because I have to rely on somebody else to bring me all the time and take me home.”

A participant mentioned finances as a barrier to participating in group programs and that having a free program was an enabler, “If this is something that helps then why wouldn’t you do it? And if it’s something that’s free and helpful, I’d be grateful for that.”

Some participants stated that the duration of the program was an enabler because it gives continuity. One participant thought 10 weeks was a good duration, “I think it’s great. It gives continuity. 10 weeks sounds like a long time but it goes in a flash.”

One participant also reported that they would stay in the program if it was well-structured with weekly goals and objectives, “I find it motivating when you have sheets to fill out, write down about how you feel and set goals every few weeks.”

#### Barriers to participation

##### Theme 4: Lack of time to follow through with the program

Several participants stated that AMD was only one of many co-morbidities that they were managing. As a result, the competing medical appointments and issues compounds difficulty in committing to the MINGLE program. An older participant who had multiple comorbidities highlighted, “I think it will be too long for me. I have a lot of doctor appointments, blood tests, exercise and other stuff…I don’t want to include anymore extra stuff.”

Participants who had to take care of other family members lacked the time to commit to the program. This was emphasised by one participant whose family member was unwell, “I think people can stay in the program when they are not busy caring for others or when they have no problem about their health…not like me because I’m caring for my husband who has so many illnesses.”

Other participants reported problems with the duration and frequency of the sessions interrupting their daily routines. Most of these participants already have sufficient social support and activities to keep them occupied and feel that accommodating 10 weeks to the program would be too long. A participant did not want the program to disturb their daily routine, “If it’s hampering or disturbing my daily routine in any big way, I wouldn’t want to be overdoing it so to say.” Another participant felt that 10 weeks would be too long to remain motivated, “I don’t know how people would get motivated…10 weeks would be too long for me. I’d get fed up with it too quickly.”

##### Theme 5: Technology as a barrier to access

Technology was mostly seen as a barrier to participation in the program. Majority of older participants who were motivated to participate saw their lack of computer or internet access as an issue. One participant stated they could not access the Internet, “I would be interested in participating…but I do have an issue with the internet access.” Another participant could not access Zoom due to lack of an appropriate device, “I don’t have access to a computer at the moment.”

However, most individuals who did not have a computer were open to trying a phone or tablet to join Zoom. Older participants with Internet or computer access mentioned that they were unable to work out how to join the online program. A participant reported that they could ask a relative or friend to help them, “I wouldn’t know how to join it, someone would need to do it for me.” Nonetheless, some were reluctant to do this because they think that it would be too burdensome, with one participant mentioning, “I’d rather do it personally, I mean I could ask my children but they are already busy as it is.”

##### Theme 6: Limited knowledge regarding the purpose of holistic interventions on quality of life

Many participants were unfamiliar with lifestyle interventions and did not understand how the program could help them. Participants with severe vision impairment believe that there is nothing that can improve their current health. This was evidenced by a participant saying, “It’s too late now because my body has already been damaged. My eyes have already been damaged. There is nothing to help.”

Other participants with mild or minimal visual impairment from AMD stated that they would prefer to spend time pursuing other activities rather than participating in the program. These participants felt that the program, particularly the social aspect, would do little to improve their current health. One participant mentioned, “In my perspective, if I had good vision, I would rather play some bowls with the guys, go to the club and throw some balls or whatever instead.” Another participant with milder AMD stated their condition was not advanced enough to experience benefit from MINGLE:

> “With macular degeneration it hasn’t affected me that much that I wouldn’t feel I would be able to discuss… people might have serious macular degeneration and I wouldn’t have anything to discuss because I’m not at an advanced stage.”

Some participants were unsure what the purpose of the program was because there were no interventions or treatments directly involving the eye. A participant asked, “Is this for the eyes or for the body? Doesn’t it look more for the legs and all that?” Another participant was confused as to how the exercises would improve their AMD, “I don’t see what the exercises do for your eyes, though.”

##### Theme 7: Vision-related issues as a barrier to participation

Severe visual impairment from AMD was also identified as a barrier to participating in the program. Since the program is delivered entirely via Zoom, loss of central vision can make it difficult for participants with advanced AMD to view the screen. A younger participant with severe visual impairment from AMD emphasised, “For some people, their vision is not like mine. Maybe they can still use the Internet or something…but for people with bad vision like me, I can’t do that.” Another participant experiencing central scotoma and distortions also echoed this sentiment:

> “I can still get by with the computer, but only just, by doing all kinds of little wriggly things to cast the light and manipulate the computer so I can get the text bigger. Yeah, so I get by, but not without a great deal of manipulating of things.”

##### Theme 8: Mobility and other health issues

Some participants viewed their lack of mobility and other health issues as a barrier to participation. Although they believed that the exercise component would be beneficial, the exercises would be difficult for them to complete due to joint pain or restriction of mobility. One participant with severe cervical radiculopathy stated, “I often have this pain so I have to lie down all day because I can’t turn my neck…I’m just worried that when it’s time for the Zoom, I won’t be well enough to join.” Another participant with chronic spinal issues also noted their inability to complete the exercises:

> “I wish I could do it… but it’s the operations… And that stops me from doing a lot of things. I’m never allowed to lift more than two kilos and I have to be careful of how I bend because of my spine.”

##### Theme 9: Negative perception of social group interactions

Some participants were disinterested in the social component of the program because others may create a negative atmosphere which deters them from engaging with others. This was highlighted by a participant who quoted, “I don’t want to socialise with people that complain about what they got. You want a positive input…we don’t want people to immerse themselves saying poor me.”

One participant mentioned that seeing other people with impaired health would cause them to feel depressed and lose confidence, “Instead of raising my self-esteem, it pulls me down…People worse than me make me feel sad and worried.”

Participants also mentioned that they did not view group socialisation, particularly online, as a crucial part of their wellbeing. A participant felt that socialising was never part of her personality, “I was never the type even when I was young, to socialise with groups and that sort of thing. It’s not me…I would just like to be left to my own thoughts.” Other participants were satisfied with their existing support networks and did not feel the need for additional socialising, as evidenced by a different participant stating, “I don’t want friends online…I have very good friends. And my children, they are good.” Another participant (89F) was concerned that individuals with dominant personalities would take over the discussion and prevent others from contributing, “I’m not a great one for groups. I tend to keep very quiet in groups…there’s always one person that seems to monopolise and you think, "Oh for goodness’ sake, sit down.”

## Discussion

This study explored the barriers and enablers to participating in the proposed MINGLE program in patients with AMD. The barriers to participation are multifactorial and include physical, psychological, social and functional factors. A multidimensional program such as MINGLE is expected to have several potential barriers as it attempts to address many facets of the biopsychosocial model simultaneously (Barrow et al., 2018). The participant responses from this qualitative study highlights the ways in which the MINGLE program can be adapted to maximise its acceptability, feasibility and efficacy.

Lack of time was a frequently reported barrier for participation. The main reasons cited in our study were concurrent health issues leading to multiple appointments, family responsibilities and reluctance to disrupt daily routines. Time-related barriers are common among other wellbeing programs for older adults (Costello et al., 2011; Kadariya & Aro, 2018). Limiting the number of sessions would most likely make the program more appealing and accessible as those with family responsibilities and appointments may be able to join. In response, the research team is currently exploring opportunities to condense the program to reduce the time-related barrier.

Many participants expressed concerns about technology and the lack of computer or Internet access. We suspect that this may be because most of our participants are of low socioeconomic status and therefore, they may not have access to the resources or education to comfortably use technology. In light of increasing adopting of information and communication technology among older adults (Australian Communications and Media Authority, 2021), especially as a result of COVID-19 and the wider integration of telehealth for the delivery of healthcare services (Garfan et al., 2021), adequate mentoring and training has been shown to overcome this barrier among adults over 65 years (Australian Government, 2018). The research has also shown that older people who are given adequate support, guidance and training when using digital interventions are more likely to develop a positive connection to them (Wilson et al., 2021). Therefore, MINGLE facilitators will offer training and education on how to use Zoom to participants before the start of the program and have an additional staff on standby during each session for technical support. Moreover, a post-intervention evaluation will be conducted with participants to assess the feasibility of in-person program delivery to bypass technology-related barriers.

Limited knowledge about the purpose of non-pharmacological lifestyle interventions was another barrier to participating in the program. Some participants with early AMD believed that the program was not targeted towards them. While some with moderate to advanced AMD felt that the program would not improve their health since it does not restore sight. However, research shows that improving lifestyle risk factors is important for people at all stages of AMD to not only reduce the risk of AMD development and its progression, but to improve overall mental and physical health and quality of life (Gastaldello et al., 2022; McGuinness et al., 2017). A lack of understanding of lifestyle risk factors is usually caused by poor health literacy and communication between patients and eyecare professionals. Despite less than one-third of adults aged 50-74 in Australia having adequate health literacy skills (Australian Institute of Health and Welfare, 2018), eyecare professionals are not providing AMD patients with the type of information and support needed to better understand their condition and how to manage it (Jalbert et al., 2020). Patient understanding of AMD risk factor modification and methods to prevent further progression such as blood pressure control, diet, antioxidant intake and cessation of smoking remains very low (Chatziralli et al., 2017; Kandula et al., 2010). Efforts to improve the awareness of AMD lifestyle risk factors is needed to address this knowledge barrier to participation in lifestyle and wellbeing interventions such as the MINGLE program.

Mobility and health concerns were a common barrier to the physical component of the program. This is consistent with existing research which states that poorer health is strongly correlated with a decrease in physical activity (Gadowski et al., 2021; Moschny et al., 2011). Older adults who have limited engagement in exercise also lack confidence in their abilities, are uncertain about the safety of physical activity and have a greater fear of injury or pain from exercise. Some older adults perceive physical activity as being unnecessary and even harmful (Franco et al., 2015). Hence, the MINGLE program will be facilitated by an Accredited Exercise Physiologist who will tailor the exercises for each participant based on their current health status so that participants can experience the benefits of physical activity without being limited by pain (Tang et al., 2022). This has been shown to increase older adults’ confidence and facilitate a more positive attitude towards physical activity (Conn et al., 2003; Franco et al., 2015).

For many participants, the main motivations to participate in the MINGLE program were the opportunity for social interaction, education and performing activities together. The social aspect of the program is important as it allows participants to interact with and motivate others who have the same condition. Social interaction is protective against social isolation and loneliness, which is a significant problem in older adults with AMD (Abdi et al., 2019; A. Brunes et al., 2019). Loneliness is a strong risk factor for poor physical and mental health and leads to premature mortality (Rokach et al., 2021). Retaining social bonds after a group intervention also increases motivation to maintain behavioural changes such as dietary and exercise habit improvements after the completion of the study (Dismore et al., 2020; Franco et al., 2015). However, not all participants in our group were receptive to social interaction. Socialisation is more likely to be accepted by participants who are older than 75 years, mostly housebound and reside alone (Kharicha et al., 2017). Due to the negative experiences by some of our participants who have previously engaged in social programs, it is critical that there is an experienced group leader to manage the discussion and participants. Therefore, for the MINGLE program, there will be a code of conduct which outlines the expected behaviour and actions of all participants to promote a safe and welcoming environment for all. Program facilitators will also make specific efforts to acknowledge and minimise the potential issues associated with group discussions including dominating personalities or overly negative talk.

Most participants stated that education was an important component of the program. Many participants would join the MINGLE program to acquire further knowledge about AMD and rationale behind its treatment, as well as the importance of improving dietary and exercise habits. Eyecare professionals have emphasised lack of patient education as one of the most significant barriers to good AMD care (Jalbert et al., 2020). This was observed in our study as some participants felt that they have limited knowledge about AMD and that there was insufficient time during consultations to ask questions. Over 50% of individuals with AMD stated that they do not receive sufficient information about diet and exercise (Stevens et al., 2014). Thus, a key benefit of the MINGLE program is that it provides opportunities to improve their understanding of AMD, its treatment and factors for progression.

### Strengths and limitations

There were several strengths in this study. Firstly, the use of semi-structured interviews allowed for tailored discussions and a more thorough understanding of the barriers and enablers to participation in the MINGLE program. We were able to capture the perspectives of older adults with AMD and identify a wide variety of themes as data was collected until saturation. There was a relatively even distribution of gender and a range of ages and ethnic groups were represented which provided a greater insight into different cultural perspectives.

The main limitation of the study was that participants were mainly recruited from a single private retinal clinic in Sydney, Australia which may limit generalisability. Most of the patients presenting to this clinic had late AMD in at least one eye and hence their opinions may differ to those with earlier stages of AMD. In addition, most of our participants are not required to pay out-of-pocket costs for their visit or treatments and would be more representative of public hospital patients rather than the majority of private retinal clinics where patients are required to pay out-of-pocket fees. Another limitation was that participant characteristics such as education levels, income and other chronic diseases that may have influenced interview responses were not collected. However, following this qualitative study, we will conduct and report on the pilot trial of the MINGLE program where this information will be collected and reported on to understand contextual factors that may influence the participants experience after completing the program.

## Conclusion

The aim of this qualitative study was to explore perceived barriers and enablers for older adults with AMD to participate in a 10-week MINGLE lifestyle program. Enablers to participation were the inclusion of non-technical information about diet, exercise and AMD, the group exercise program and motivation to improve patient health. Barriers to participation were primarily due to the lack of time, patient understanding about wellbeing programs and difficulties with digital access. All these factors will be taken into consideration during the development and implementation of the MINGLE program. More efforts are needed to raise patient awareness of non-pharmacological treatment of AMD.

## Data Availability

All data produced in the present study are available upon reasonable request to the authors

