## Appendix 1 and 2 for "Barriers and enablers to participation in a proposed online lifestyle intervention for older adults with age-related macular degeneration"

Appendix 1. Interview Guide

| **Area of Discussion** | **Questions and Prompts** |
| --- | --- |
| Icebreaker | Briefly describe yourself and what keeps you busy?  What kind of AMD do you have and how has it affected your day-to-day life? |
| Previous experience with social programs | Have you ever participated in a social program for seniors? If so, what did the program involve and how did you find the program? |
| MINGLE intervention design | What are your thoughts on the duration and frequency of the sessions? What would your preference be in terms of day and time if you were to participate in the MINGLE program?  What are your thoughts on the exercise and nutrition topics? Do you have suggestions on what to include/exclude in the socialising component? If so, why?  Do you think that there is anything else that should be included or excluded from the MINGLE program? If so, why? |
| Motivation | Could you describe any concerns you have about participating in the MINGLE program?  What do you think might be the best way to keep participants motivated and committed to the 10-week MINGLE program?  Do you have any further comments regarding the MINGLE program or anything else that we have spoken about today? |

Appendix 2. Additional Interview Quotes

| **Construct** | **Themes** | **Quotes** |
| --- | --- | --- |
| Enablers to participation | Opportunity to meet new people and learn more about AMD | “We need that, especially me and my husband, because the whole day, the whole night, it’s only just me and my husband. So, we need to socialise.” |
|  |  | “I'm 86, I still live a very active life. I play bowls three times a week. I live in a community centre and we do stuff with all our people. You know, we have bingo and all that. We get together very often. You know, we have lunches and we have outings.” |
|  |  | “You could talk about your condition, you can talk about politics or about any current events.” |
|  |  | “There's one big topic I think that should be brought in really plain language that elderly people can understand. And that is why they need the injections, what it's doing and, and the goodness it's doing to them, because there's a lot of them just say, "Well, my doctor told me, I'll have to have it done. And I don’t know why." And I mean, if you could just introduce just something in plain English brought down so that they can understand and just say, "Well, right. It's because you have macular degeneration and it does this, and it does that, and the injections will help you."” |
|  |  | “There's a lot of elderly people…here I am at 84 and we just don't have any knowledge of what's happening.” |
|  |  | “I live on my own and you need to socialise when you're on your own. COVID was a bit of a challenge.” |
|  | Participants expressed motivation to improve their current health and the health of others | “It's a pretty serious business when it comes to your eyes. Nobody wants to go blind.” |
|  |  | “Appealing to the sense of helping future people, because five years down the track, it could be somebody else they know and if they can help them, then all good.” |
| Barriers to participation | Lack of time to follow through with the program | “As long as it doesn’t interfere with appointments or anything, it doesn’t matter what time it’s done” |
|  |  | “Maybe for younger people it’s better, but for me I’ve already got enough. Why would I want to do more hours like that, my plate is already full.” |
|  |  | “If it’s hampering or disturbing my daily routine in any big way, I wouldn’t want to be overdoing it so to say.” |
|  | Limited knowledge regarding AMD lifestyle risk factors | “My mother, she's passed away now, but she had macular degeneration. It virtually sent her blind and she ended up living in a nursing home the last couple of years of her life. But, uh, yeah, she had, she had big problems with it because they had very limited medical procedures to help in that regard.” |
|  | Vision-related issues as a barrier to participation | “You see like when I’m watching TV I can’t see properly. I see lots of shadow pictures, you know? Before when I had good eyes, I joined everything, you know? My eyes are not working as good now so I can't join that anymore.” |
|  |  | “I used to do a lot of things, but the eye situation has stopped me from doing many of them. I used to race cars and motorbikes and boats. Flying airplanes. But all those things I can't do anymore.” |
|  | Mobility and other health issues | “I couldn’t join because my eye, my body and spinal pain…the movements, it’d be very, very difficult to do those.” |
|  |  | “If I was 60, 70, I would recommend everybody to do this. But now one day I am okay and the next day I am run down. I don’t have the energy to be on the program all the time.” |
|  |  | “With the exercises, my muscles are already stiff. If I do exercises, it's painful for me because I need massage more than exercises. When I do exercises, I’m always in pain. The more I'm doing, the more pain I have…I tell you the truth, in one hour the only thing I wanted is to rest.” |
|  |  | “I have lung cancer at the moment and I just buckled my way through bowel cancer. I’ve got a couple of little polyps in the lung so I’m starting chemotherapy and I really can't join. I have it once a fortnight and by the time I get around to having it again, I've just about got over the first. So really, at the moment I can't do it, but I mean, I would like to do it later on after I get rid of this.” |
|  | Lack of interest or negative perception of social group interactions | “This one guy here, oh my god. He went on and on and on about all the things were wrong with him and I'm thinking to myself, you're not going to get better anyways if all you're focusing on is you know...On your demise. It's just like, go, just dig a hole, darling. Go and lay down and put your arms right like that and say, 'goodbye everybody'. Because that's where you're going to end up, you know.” |
|  |  | “Well, to be honest, I never speak about my problems.” |
|  |  | “Sometimes it’s not a pleasant experience…so I can either go into it and go, “Yeah, I don’t like it” or I can go in with a positive mindset and set myself up for a positive session.” |
|  |  | “Just time commitments and things like that, you know what I mean? It’s not my cup of tea. I’ve got things to do…me and my wife we do all sorts of stuff.” |
